# Association between COVID-19 Risk-Mitigation Behaviors and Specific Mental Disorders in Youth

**DOI:** 10.1101/2022.03.03.22271787

**Authors:** Kevin P. Conway, Kriti Bhardwaj, Emmanuella Michel, Diana Paksarian, Aki Nikolaidis, Minji Kang, Kathleen R. Merikangas, Michael P. Milham

## Abstract

**Importance:** Although studies of adults show that pre-existing mental disorders increase risk for COVID–19 infection and severity, there is limited information about this association among youth. Mental disorders in general as well as specific types of disorders may influence their ability to comply with risk-mitigation strategies to reduce COVID-19 infection and transmission.

**Objective:** To examine associations between specific mental disorders and COVID-19 risk-mitigation practices among 314 female and 514 male youth.

**Design:** Youth compliance (rated as “Never,” “Sometimes,” “Often,” or “Very often/Always”) with risk mitigation was reported by parents on the CoRonavIruS Health Impact Survey (CRISIS) in January 2021. Responses were summarized using factor analysis of risk mitigation, and their associations with lifetime mental disorders (assessed via structured diagnostic interviews) were identified with linear regression analyses (adjusted for covariates). All analyses used R Project for Statistical Computing for Mac (v.4.0.5).

**Setting:** The Healthy Brain Network (HBN) in New York City Participants. 314 female and 514 male youth (ages 5-21)

**Main Outcome(s) and Measure(s):** COVID-19 risk mitigation behaviors among youth

**Results:** A two-factor model was the best-fitting solution. Factor 1 (avoidance behaviors) included avoiding groups, indoor settings, and other peoples’ homes; avoidance was more likely among youth with any anxiety disorder (p=.01). Factor 2 (hygiene behaviors) included using hand sanitizer, washing hands, and maintaining social distance; practicing hygiene was less likely among youth with ADHD (combined type) (p=.02). Mask wearing, which did not load on either factor, was not associated with any mental health disorder.

**Conclusion and Relevance:** Findings suggest that education and monitoring of risk-mitigation strategies in certain subgroups of youth may reduce risk of exposure to COVID-19 and other contagious diseases. Additionally, they highlight the need for greater attention to vaccine prioritization for individuals with ADHD.

**Key Points:** *Question:* Are mental disorders among youth associated with COVID-19 risk-mitigation behaviors?

*Findings:* Based on the parent CoRonavIruS Health Impact Survey (CRISIS) of 314 females and 514 males aged 5-21, youth with anxiety disorders were more likely to avoid high-risk exposure settings, and those with ADHD (combined type) were less likely to follow hygiene practices. In contrast, mask wearing was not associated with youth mental disorders.

*Meaning:* Specific types of disorders in youth may interfere with their ability to employ risk-mitigation strategies that may lead to greater susceptibility to COVID-19.

## Introduction

The COVID-19 pandemic constitutes an ongoing global public health threat. SARS-CoV-2 is a highly contagious virus that transmits mainly through inhalation of airborne droplets and transfer from direct contact with surfaces that are contaminated. Public health officials across the world initially responded to this threat by urging people to follow several risk-mitigation strategies to reduce the chance of infection and transmission. Adults and children alike were urged to avoid close contact, maintain at least 6-feet of physical distance from others, wear face masks in public, engage in frequent and intensive hand washing, adhere to stay-at-home orders, and self-isolate when exhibiting symptoms of infection^1^. Such efforts have been associated with a reduction in COVID-19 transmission^2-4^.

There is growing evidence regarding the potential impact of mental health disorders on COVID-19 risk. Studies of adults show that pre-existing mental disorders are associated with increased risk of COVID-19 infection^5^, severity^6-9^, and mortality^7-12^. A recent study showed that individuals with substance use disorders are also at increased risk of COVID-19 infection, hospitalization, and death^8,9^. These findings have been largely confirmed by several meta-analyses^7-9,13^.

Accordingly, the Centers for Disease Control designated mental and behavioral health disorders among the pre-existing conditions that increase vulnerability to COVID-19 illness. An important effect of this recognition is greater priority in receiving vaccines and boosters as they are rolled out, which help to protect individuals and reduce community spread. Included in this list are disorders that typically affect children, such as neurodevelopmental disorders like autism spectrum disorder and ADHD. However, there is scant research on the role of mental disorders in COVID-19 vulnerability among youth. One recent study of electronic health records among patients (aged 2 months to 103 years) in Israel found that COVID-19 infection was associated with ADHD (but no other psychiatric diagnosis examined), male gender, age below 20 years, and low-medium SES group^14^. Interestingly, the association between ADHD and COVID-19 infection in this sample was especially elevated among youth (ages 5-20) and untreated ADHD cases of any age. Additional research on youth with a range of mental disorders is needed to understand the potential mechanisms associated with COVID-19 vulnerability. In particular, the ability of youth to adhere to risk-mitigating practices may differ by the key phenomena underlying mental disorders such as inattention, anxiety, fear, impulsivity, etc. To our knowledge, no studies have investigated COVID-19 risk-mitigation adherence among youth with mental disorders. The purpose of this study was to examine associations between COVID-19 risk-mitigation practices and specific mental disorders among female and male youth from the Healthy Brain Network (HBN) in the New York City metropolitan area.

## Methods

### Sample

Participants were recruited from the ongoing Healthy Brain Network (HBN) initiative that seeks to create and share a 10,000-participant biobank of data from children and adolescents ages 5–21 from the New York City area^15^. Data collected includes psychiatric, cognitive, behavioral, genetic, and lifestyle information as well as MRI and EEG neuroimaging. The HBN collection sites are on Staten Island, in Midtown Manhattan, and in Harlem. As part of the HBN survey battery, participants and their parents/guardians completed a variety of age-based questionnaires assessing basic demographic characteristics, dimensional assessments of domains associated with mental health, substance use, and socioeconomic status. For participants under the age of 11, a trained research assistant read and explained individual items and collected responses from participants. HBN’s latest data release includes 4139 participants; data are available to researchers by registering for a data usage agreement (http://fcon_1000.projects.nitrc.org/indi/cmi_healthy_brain_network/Pheno_Access.html#DUA).

Between April and July 2020 (Wave 1), parents of HBN participants were invited to complete the CoRonavIruS Health Impact Survey (CRISIS)^16^ about their child via Research Electronic Data Capture (REDCap). The CRISIS was designed and piloted at the beginning of the COVID-19 pandemic to assess mental and behavioral health, lifestyle behaviors, and sources of stress induced by the COVID-19 epidemic. In total, parents of 1780 HBN participants completed the Wave 1 survey. Parents were then invited to complete a modified version of the CRISIS in January 2021 (Wave 2). The Wave 2 modifications included questions on frequency of compliance with COVID-19 risk-mitigation practices. The current study sample included 314 female and 514 male participants (ages 5-21) whose parents completed the CRISIS at Wave 2.

### Measures

Measures derived from HBN participation included child age at Wave 2, sex, race/ethnicity, family structure, family socioeconomic status (SES), and consensus diagnosis. Child age, sex, race/ethnicity (Caucasian, African American, Hispanic, Asian), and family structure (indicator of single caregiver household) were reported by parents/caregivers during a structured clinical history interview. Family SES was measured by the Barratt Simplified Measure of Social Status, which is based on parent/caregiver reports of parent/caregiver education and occupation^17^. Continuous scores are generated with higher scores indicating higher SES. In single caregiver families, scores were based on that caregiver alone. SES scores were subsequently grouped into tertiles to determine low, middle, and high SES.

Diagnostic interviews were conducted using the computerized Kiddie Schedule for Affective Disorders and Schizophrenia^18^ that was administered to parents by an experienced research clinician or social worker. Following Diagnostic and Statistical Manual of Mental Disorders, Fifth Edition (DSM-5) criteria, consensus lifetime diagnosis was achieved by two study psychiatrists based on these interviews and other symptomatic information such as standardized rating scales. Up to 10 separate diagnoses were allowed per participant. Diagnoses were grouped into the following categories: attention deficit hyperactivity disorder inattentive/hyperactive type [ADHD-I), ADHD combined type (ADHD-C), autism spectrum disorder, any depressive disorder (major depressive disorder [MDD], persistent depressive disorder [PDD], disruptive mood dysregulation disorder [DMDD], depressive disorder due to another medical condition, unspecified depressive disorder, substance/medication-induced depressive disorder, other specified depressive disorder, other (or Unknown) substance-induced disorders), any anxiety disorder (unspecified anxiety disorder, generalized anxiety disorder [GAD], separation anxiety, social anxiety, specific phobia, agoraphobia, panic disorder, selective mutism, other specified anxiety disorder), and any other behavior disorder (oppositional defiant disorder, conduct disorder, intermittent explosive disorder, or other specified disruptive, impulse-control disorder).

Seven separate COVID-19 risk-mitigation practices among youth were measured via parent report at Wave 2 of CRISIS administration. Specifically, parents were asked with respect to the past two weeks “To what extent has your child been taking the following steps to prevent infection or spread of the virus? Wearing a mask or face covering in public; Wearing gloves in public; Washing hands; Using hand sanitizer; Staying at least six feet away from others; Avoiding visits to other people’s homes; Avoiding group in-person activities; Avoiding indoor public places (e.g., stores) when possible.” Responses were rated as “Never,” “Sometimes,” “Often,” or “Very often/Always.”

### Analysis

Primary analyses were conducted using the parent CRISIS survey conducted in January 2021. This survey was limited to the 1578 past HBN participants (2015-2020; total: 3600) that completed an initial survey in April-June 2020 (Wave 1). 955 parents completed the survey in 2021. Participants that did not complete enough of the HBN study to yield a consensus DSM diagnostic profile based on the KSADS-COMP and licensed clinical evaluators, were removed. Five participants with missing Barratt data were also removed. The final analytic sample comprised 314 female and 514 male participants between ages 5-21 years (N=828). Sample characteristics did not differ by study-completion status (see Supplement). All statistical analyses were conducted using The R Project for Statistical Computing for Mac^19^.

Responses to the 7 risk-mitigation practices were summarized using factor analysis, and model fitness was evaluated by parallel analysis. Associations between the resulting factors and lifetime mental disorders were identified with linear regression analyses. Three models were used in the analysis: unadjusted, adjusted for demographics (age, sex, SES, single caregiver, and race), and additionally adjusted for comorbid mental disorders. Each factor underwent the three-model analysis to examine the associations between specific mental disorders and each derived factor of risk-mitigation behaviors.

## Results

Table 1 presents the number and percentages of youth who “very often/always” engaged in the 7 risk-mitigation behaviors, by demographic characteristics. Overall, the percentage of mask wearing “very often/always” was high (90%), and higher in females, African Americans, and Hispanics. The overall percentage of “very often/always” maintaining social distance, using hand sanitizer, and washing hands was 46%, 46%, and 59%, respectively. These behaviors differed significantly by race, as the percentages were elevated among African Americans and Hispanics and lower among Caucasians. Further, maintaining social distancing “very often/always” was positively associated with age and negatively associated with SES, as was washing hands “very often/always.” The overall percentages of avoiding other people’s homes, avoiding in-person groups, and avoiding indoor public places “very often/always” was 58%, 46%, and 43%, respectively. These were highly similar across demographic factors.

**Table 1.** Participant Demographics by COVID-19 Risk-Mitigation Items.

|  |  | COVID-19 Risk-Mitigation Behaviors <sup>1</sup> |  |  |  |  |  |  |
| --- | --- | --- | --- | --- | --- | --- | --- | --- |
|  | Total Analytic Sample | Mask Wearing | Maintaining Social Distance | Using Hand Sanitizer | Washing Hands | Avoiding Other People's Homes | Avoiding In-Person Groups | Avoiding Indoor Public Places |
| **Sex** |  | * |  |  |  |  |  |  |
| Male | 514 (62%) | 452 (88%) | 223 (43%) | 225 (44%) | 297 (58%) | 307 (60%) | 228 (44%) | 224 (44%) |
| Female | 314 (38%) | 293 (93%) | 149 (47%) | 159 (51%) | 195 (62%) | 174 (55%) | 150 (48%) | 136 (43%) |
| **Age (years)** |  |  | * |  |  |  |  |  |
| 5-6 | 20 (2%) | 19 (95%) | 6 (30%) | 10 (50%) | 16 (80%) | 13 (65%) | 6 (30%) | 9 (45%) |
| 7-9 | 217 (26%) | 193 (89%) | 85 (39%) | 105 (48%) | 136 (63%) | 135 (62%) | 98 (45%) | 96 (44%) |
| 10-12 | 302 (36%) | 273 (90%) | 134 (44%) | 133 (44%) | 171 (57%) | 162 (54%) | 132 (44%) | 130 (43%) |
| 13-15 | 178 (21%) | 161 (90%) | 86 (48%) | 83 (47%) | 101 (57%) | 111 (62%) | 93 (52%) | 82 (46%) |
| 16+ | 111 (13%) | 99 (89%) | 61 (55%) | 53 (48%) | 68 (61%) | 60 (54%) | 49 (44%) | 43 (39%) |
| **Family Structure** |  |  | * |  |  |  | * |  |
| Single caregiver | 70 (8%) | 66 (94%) | 40 (57%) | 37 (53%) | 45 (64%) | 44 (63%) | 42 (60%) | 33 (47%) |
| **SES <sup>4</sup>** |  |  | * |  | * |  |  |  |
| Low | 58 (7%) | 54 (93%) | 32 (55%) | 31 (53%) | 41 (71%) | 33 (57%) | 33 (57%) | 25 (43%) |
| Middle | 153 (18%) | 138 (90%) | 80 (52%) | 77 (50%) | 102 (67%) | 83 (54%) | 76 (50%) | 65 (42%) |
| High | 617 (75%) | 553 (90%) | 260 (42%) | 276 (45%) | 349 (57%) | 365 (59%) | 269 (44%) | 270 (44%) |
| **Race** |  | * | * | *** | *** |  |  |  |
| Caucasian | 444 (54%) | 390 (88%) | 174 (39%) | 174 (39%) | 226 (51%) | 249 (56%) | 189 (43%) | 195 (44%) |
| African American | 99 (12%) | 96 (97%) | 57 (58%) | 60 (61%) | 70 (71%) | 61 (62%) | 55 (56%) | 46 (46%) |
| Hispanic | 71 (9%) | 66 (93%) | 42 (59%) | 40 (56%) | 49 (69%) | 35 (49%) | 33 (46%) | 26 (37%) |
| Asian | 27 (3%) | 24 (89%) | 11 (41%) | 12 (44%) | 17 (63%) | 18 (67%) | 10 (37%) | 12 (44%) |
| Other | 158 (19%) | 140 (89%) | 73 (46%) | 79 (50%) | 109 (69%) | 100 (63%) | 76 (48%) | 71 (45%) |
| Unknown | 29 (4%) | 29 (100%) | 15 (52%) | 19 (66%) | 21 (72%) | 18 (62%) | 15 (52%) | 10 (34%) |
| **Site<sup>5</sup>** |  |  |  |  |  |  |  |  |
| Staten Island<sup>6</sup> | 288 (35%) | 266 (86%) | 124 (43%) | 139 (48%) | 164 (57%) | 153 (53%) | 126 (44%) | 114 (40%) |
| Midtown | 210 (25%) | 197 (90%) | 101 (48%) | 89 (42%) | 123 (59%) | 124 (59%) | 102 (49%) | 100 (48%) |
| Harlem | 322 (39%) | 304 (93%) | 144 (45%) | 151 (47%) | 199 (62%) | 200 (62%) | 147 (46%) | 143 (44%) |

Note. <sup>1</sup> Defined as “Very often/Always” according to the original responses of “Never,” “Sometimes,” “Often,” or “Very often/Always.” Chi-Square group differences are represented by asterisks; \* p < 0.05, \*\* p < 0.01, \*\*\* p < 0.001. <sup>4</sup> Barratt total score was divided into tertiles: Low (3-24), medium (25-45), high (46-66). <sup>5</sup>Mobile Research Vehicle (MRV) site not shown in table (N=8, %=1). <sup>6</sup> Combined Staten Island site and Staten Island Richmond University Medical Center site.

The correlations among the risk-mitigation items appear in Figure 1. All correlations were positive and ranged from 0.09 to 0.63. Mask wearing correlated weakly (r<0.31) with all other items. Maintaining social distance correlated moderately (*r*=0.38 to *r*=0.44) with using hand sanitizer, avoiding other people’s homes, avoiding in-person groups, and avoiding indoor public places, and washing hands. Using hand sanitizer correlated strongly (*r*>0.63) with washing hands and weakly (*r*<0.14) with avoiding other people’s homes, avoiding in-person groups, and avoiding indoor public places. Washing hands correlated weakly (*r*<0.21) with avoiding other people’s homes, avoiding in-person groups, and avoiding indoor public places. The three avoidance items – avoiding other people’s homes, avoiding in-person groups, and avoiding indoor public places – were moderately to strongly intercorrelated (*r*=0.53 to *r*=0.60).

**Figure 1.**
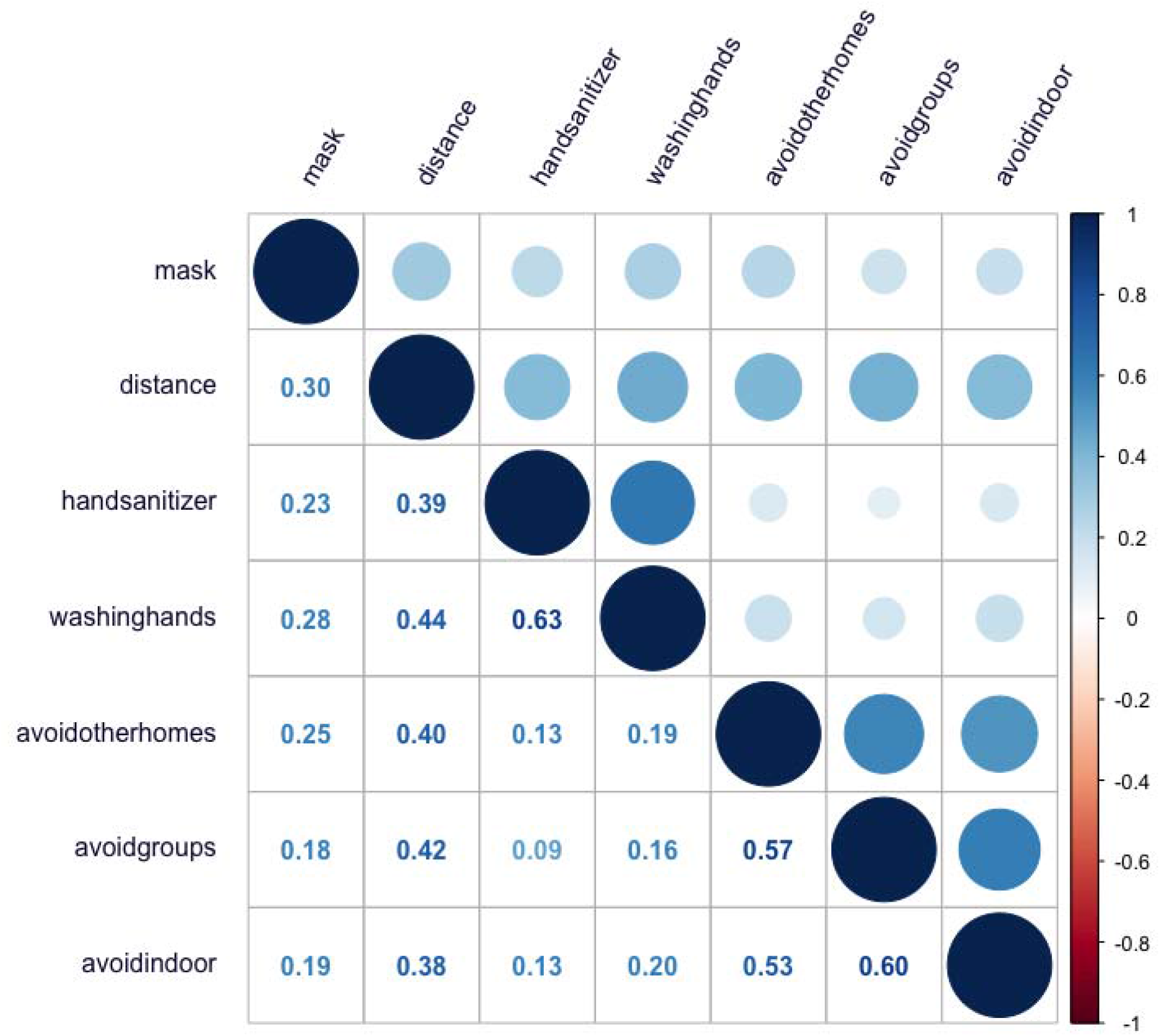
Correlation Matrix for COVID-19 Risk-Mitigation Items.

Results from the parallel analysis (Figure 2) show that a two-factor model was the best-fitting solution. That is, a two-factor solution explained more variance than what would be expected due to chance based on a null distribution of eigenvalues. Results from the factor analyses are shown in Figure 3. The first factor (avoidance behaviors) included avoiding groups, indoor settings, and other peoples’ homes. This factor accounted for 27.7% of the cumulative variance. The second factor (hygiene behaviors) included using hand sanitizer, washing hands, and maintaining social distance. The addition of this factor increased the cumulative variance explained to 49.8%. Mask wearing loaded poorly on each factor, a finding consistent with the weak individual correlations with this item. Further, its inclusion to the model did not meaningfully improve the fit or the amount of variance explained. For these reasons, mask wearing was analyzed subsequently as a separate item.

**Figure 2.**
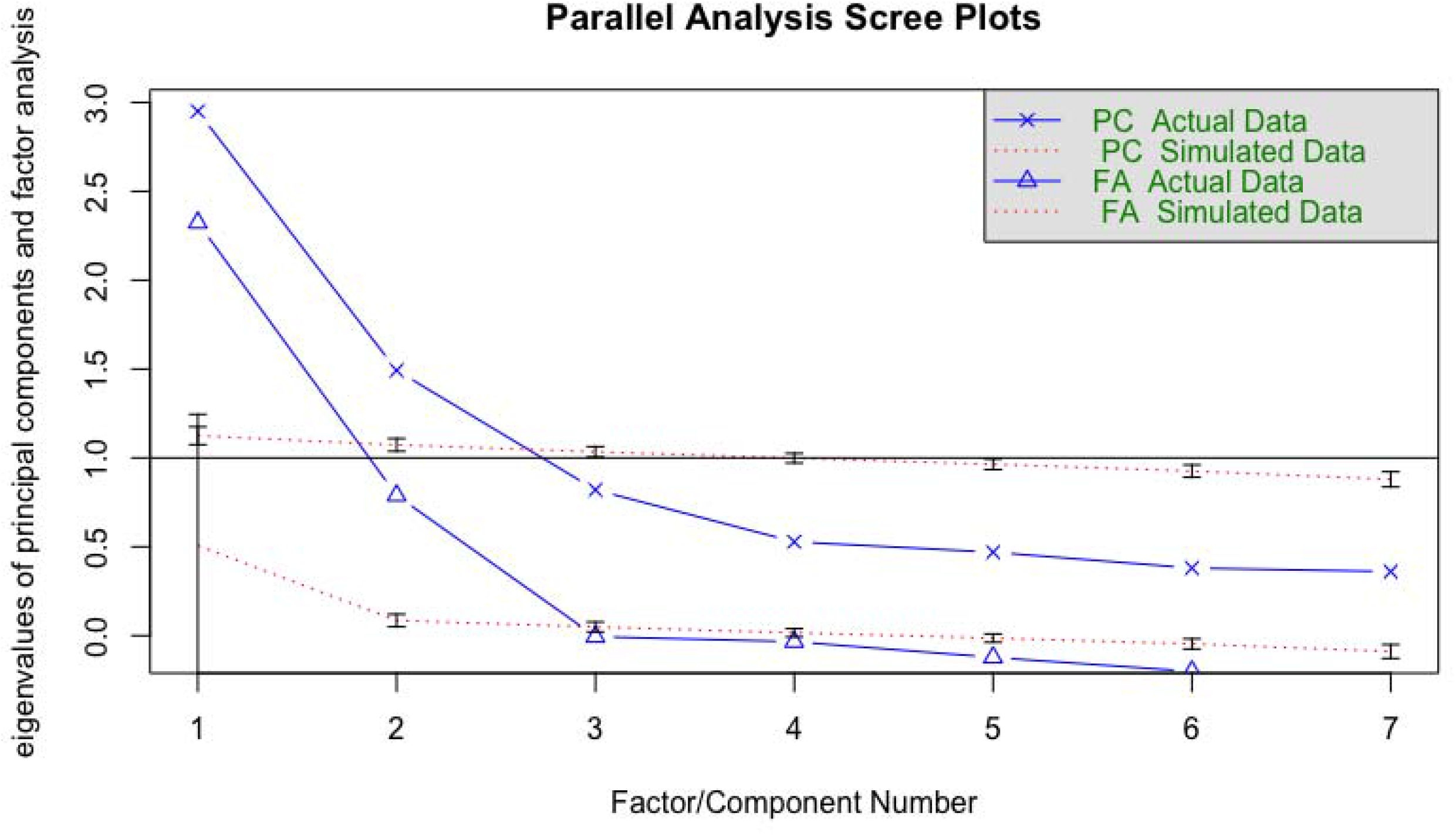
COVID-19 Risk-Mitigation: Results from Parallel Analysis.

**Figure 3.**
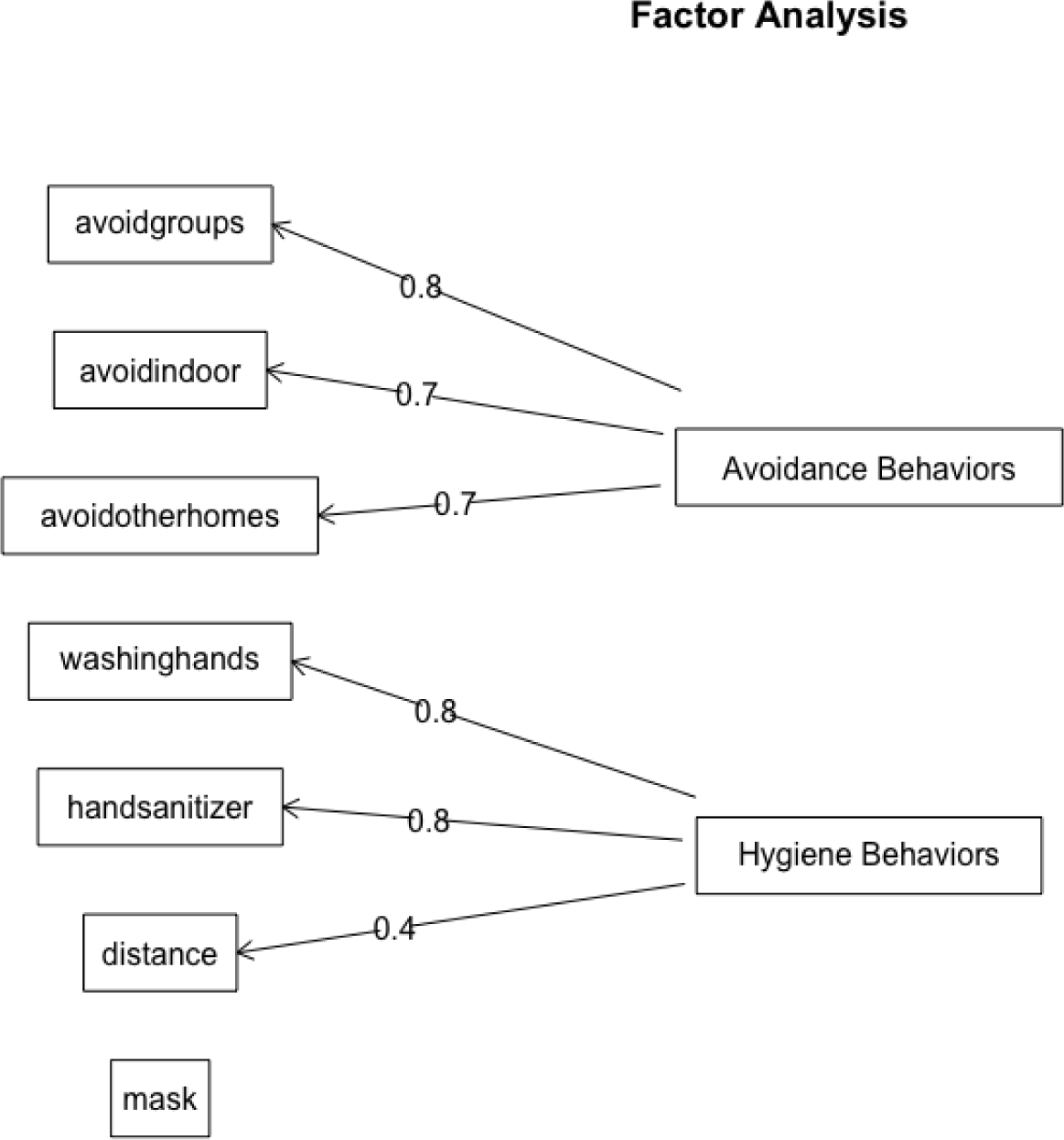
COVID-19 Risk-Mitigation: Results from Factor Analysis.

Table 2 presents the results from the regression analyses examining associations between the two resulting factors (and, separately, mask wearing) and mental disorders. The table presents results from the unadjusted model, the model (a) adjusted for demographics, and the model (b) adjusted for demographic variables and comorbid mental disorders. Focusing on the fully adjusted model (b), several of the demographic variables were significant covariates (results not shown). African American race was positively associated with both avoidance behaviors and hygiene behaviors. Age, Hispanic race, other race, or nondisclosed race were positively associated with hygiene behaviors. Male sex was positively associated with mask wearing. As shown in Table 2, the two factors were associated with specific mental disorders. The avoidance behaviors were more likely among youth with any anxiety disorder, a finding that was significant in the unadjusted model (*p*=.008), the model that adjusted for demographic variables (*p*=.01), and the model that adjusted for demographics and comorbid disorders (*p*=.01). No other disorder was significantly associated with this factor. The hygiene behaviors were less likely among youth with ADHD-C in the unadjusted model (*p*=.01), the model that adjusted for demographic variables (*p*=.02), and the model that adjusted for demographics and comorbid disorders (*p*=.02). Hygiene behaviors were negatively associated with depression, but only in the unadjusted model (*p*=.04). Analyzed as a separate item, mask wearing was less likely among youth with ADHD-C in the unadjusted model (*p*=.03). However, this association was not significant after adjusting for demographics and comorbid disorders. No other mental disorder was significantly associated with mask wearing.

**Table 2.**
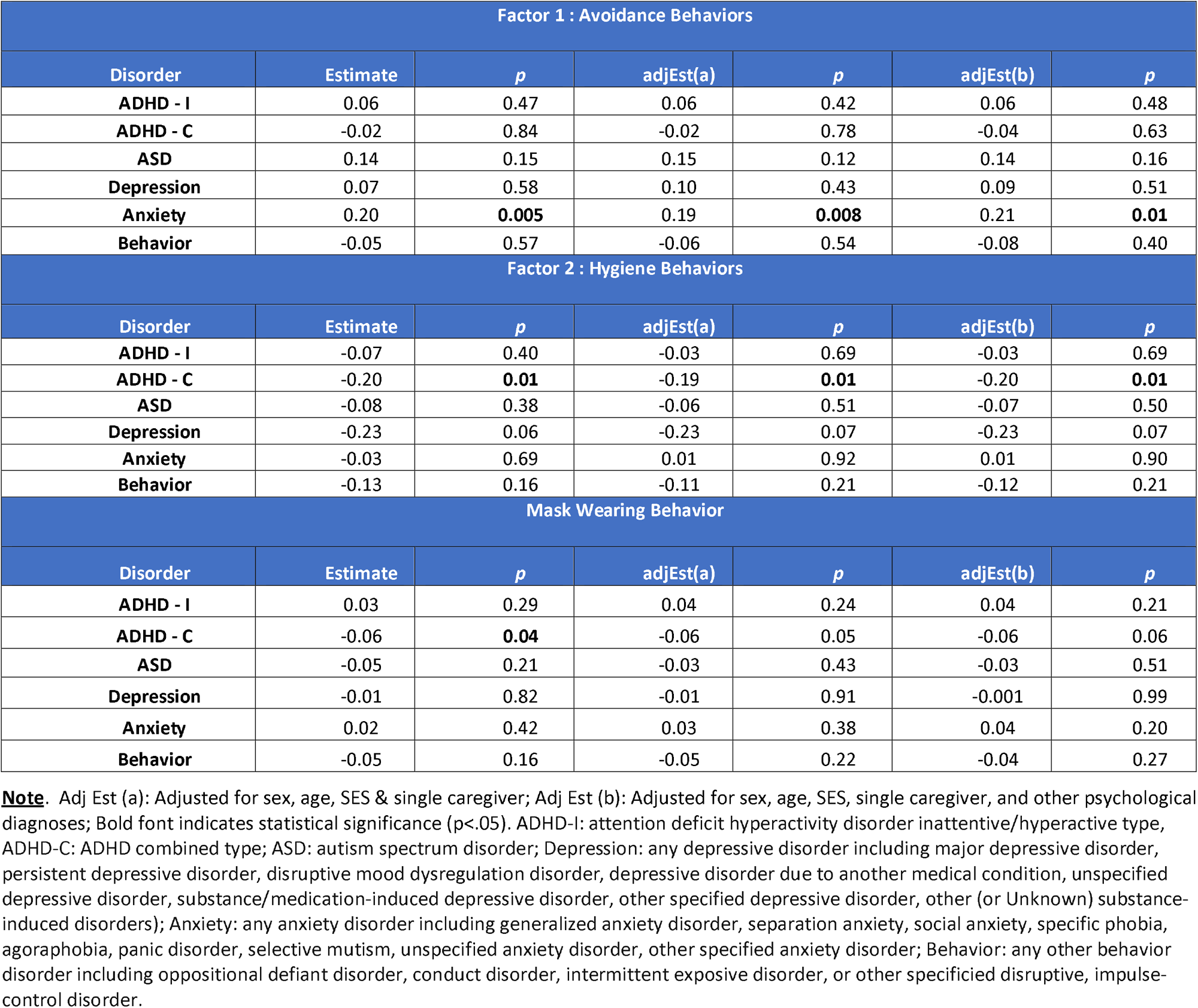
Associations between Risk-Mitigation Behaviors and Lifetime Psychiatric Disorders.

## Discussion

The importance of risk-mitigation measures in reducing exposure and severe disease have been one of the most important public health measures in response to the prolonged nature of the COVID-19 pandemic, now entering its third year. Physical distancing, avoiding touch, washing hands frequently and intensively, wearing a face mask in public, staying at home, and maintaining quarantine have been particularly challenging for youth with mental disorders. Here we report two important findings regarding the association between compliance with different risk-mitigation factors with different types of disorders. First, the COVID-19 risk-mitigation behaviors tended to cluster into two factors: avoidance behaviors (avoiding groups, indoor settings, and other peoples’ homes) and hygiene behaviors (using hand sanitizer, washing hands, and maintaining social distance). Second, the tendency to practice these behaviors differed by specific mental disorders. Avoidance was more likely among youth with any anxiety disorder, whereas practicing hygiene was less likely among youth with ADHD-C. Mask wearing was largely unrelated to other risk-mitigation behaviors and, when examined as a separate item, was not associated with mental disorders after adjusting for demographic factors or comorbid mental disorders.

Our findings add to the limited research on the role of mental disorders in COVID-19 risk among youth, despite ample evidence showing that children and adolescents with mental disorders are at increased risk of myriad physical diseases^20^. In particular, ADHD has been shown increase risk of inflammatory and immune-related disorders including asthma, eczema, certain allergies^21-24^, respiratory infections and influenza^12^, and several preventable negative outcomes including sexually transmitted infections, accidental injuries, and injury-related mortality^25-27^. Psychosocial stress, anxiety, negative affect, and depression are also associated with increased risk of acute respiratory infections as well as poorer clinical prognosis^28^. Depression has been associated with an increased risk of a developing a wide range of infections^29^ including sexually transmitted diseases and poorer outcomes thereof^30,31^. Children and adolescents with autism spectrum disorders are at increased risk of several medical conditions including immunological, gastroenterological, neurological, and other medical complaints^22^. This work provides some insight into the increased risk by showing that youth with specific mental disorders are more (or less) likely to engage in certain COVID-19 risk-mitigation behaviors.

There are several potential mechanisms for our findings. Youth with anxiety disorders, who were more likely to practice avoidance behaviors, may be so inclined because avoidance is a central characteristic of many anxiety disorders including social phobia, agoraphobia, social anxiety disorder, and generalized anxiety disorder. Avoidance-related manifestations of anxiety disorder may serve to mitigate COVID-19 vulnerability by reducing social contacts with others in groups, indoor settings, and other peoples’ homes. The failure to comply with hygiene behaviors among youth with ADHD-C may occur through other mechanisms. Youth with ADHD-C, where both inattention and hyperactivity/impulsivity are present, may be less capable of complying with 6-foot distance rules and hand-cleaning practices. This may help explain the finding reported by Merzon and colleagues^14^ that youth with ADHD are at increased risk of COVID-19 infection, and that the association was greater for untreated than treated ADHD. Several non-significant findings are notable as well. Although youth with ASD can sometimes struggle with daily living skills including maintaining personal hygiene^32^ that may lead to lower compliance with hand washing and using hand sanitizer, we did not observe this finding. Youth with oppositional defiant disorder or conduct disorder, who tend to disregard social rules, were no more or less likely to consistently engage in harm-mitigation behaviors. We similarly found no independent association between depressive disorders and risk-mitigation behaviors. In summary, these interpretations are suggestive and warrant further research.

Aside from risk-mitigation compliance, there are other mechanisms by which pre-existing mental and behavioral health disorders may increase COVID-19 vulnerability. These include increased exposure to COVID-19 at home or at school due to greater household or community density, greater susceptibility due to enhanced physical disease vulnerability as described above, or poorer general health status related to obesity, physical inactivity, or familial exposure to smoking. Future research on COVID-19 vulnerability should incorporate the full range of risk factors for COVID-19 infection as well as compliance with COVID-19 risk-mitigation behaviors.

To further reduce COVID-19 incidence and community transmission, it is crucial to identify risk factors for COVID infection in youth. Although some research shows that young children are less susceptible to infection than adults^33-35^, other research shows that children may be as likely as adults to become infected with COVID-19^36^. In fact, a recent epidemiologic study^37^ of a pediatric sample in Virginia reported a SARS-CoV-2 infection rate (8.5%) that was higher than a sample of adults (2.4%) from a similar region and period^38^. Further, children appear to play a role in community transmission through their social interactions and hygienic habits^33^, a finding that underscores the importance of risk-mitigation strategies among youth especially as the COVID-19 pandemic continues to evolve and become even more transmissible, as in the case of the most recent circulating variant (Omicron). Indeed, a recent experiment found that the Omicron variant survives longer than other variants on plastic and skin, a factor that may have contributed to the rapid community spread of Omicron^39^. Taken together, findings further underscore the importance of being vigilant about risk-mitigation behaviors to combat vulnerability to COVID-19 infection and transmission.

This work provides novel information on the associations between mental disorders and COVID-19 risk-mitigation behaviors in youth. Reduced practice of prevention measures among those with specific types of disorders highlights the need to gain greater insight into the specific difficulties underlying the reduced compliance observed here. Risk-mitigation education could then be tailored to the specific components of these conditions that reduce compliance efforts. It may also be worthwhile to consider prioritizing vaccinations among individuals with mental and neurodevelopmental disorders^40^ that may reduce their ability to prevent exposure.

## Data Availability

All data produced in the present study are available upon reasonable request to the authors.

---

**Supplement.**
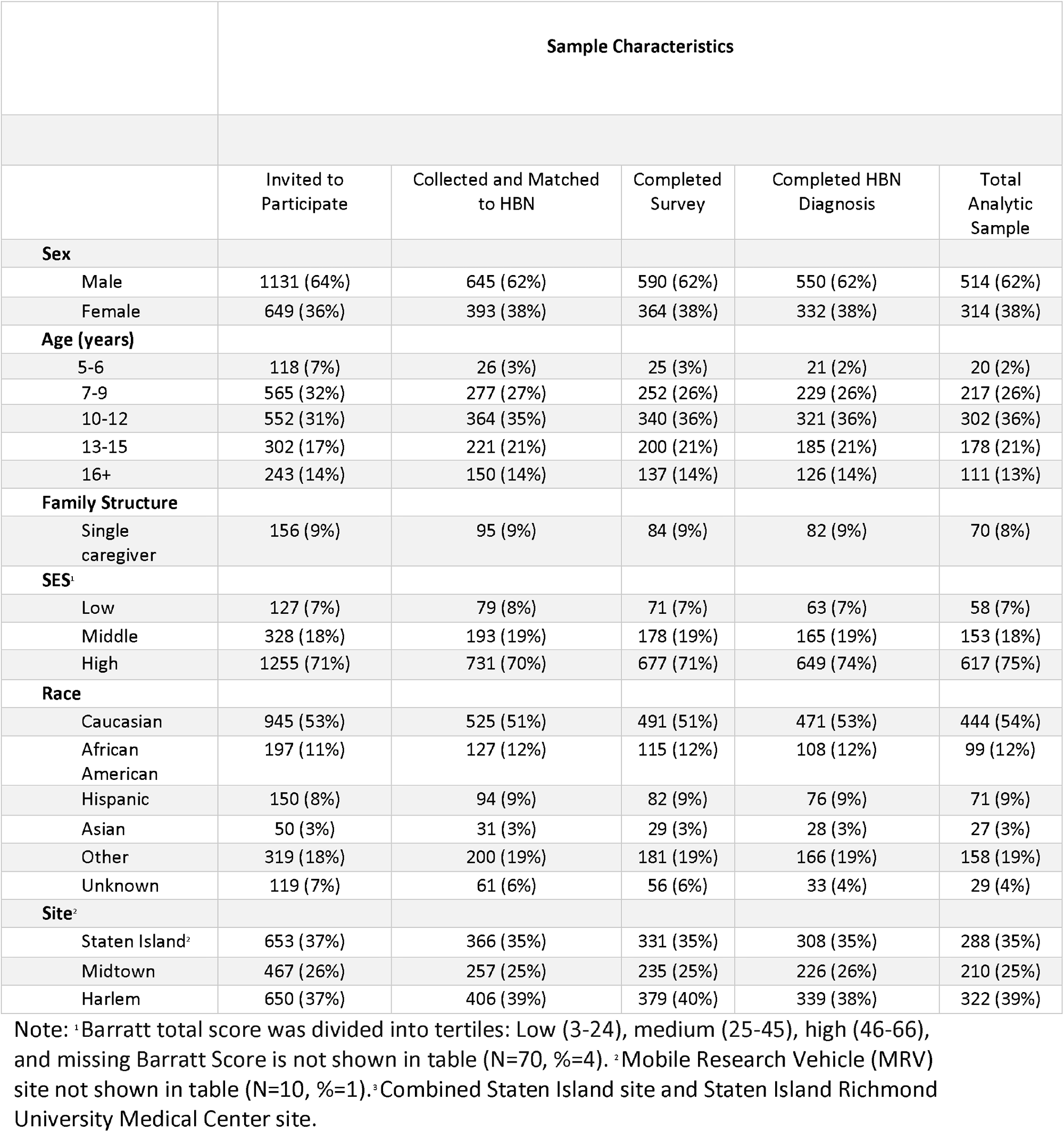
Sample Characteristics by Study Completion Status.

## Notes

Funding: This work was supported in part by the Intramural Research Program of the National Institute of Mental Health (ZIAMH002953) and Morgan Stanley. Additionally, by philanthropic gifts from Phyllis Green, Randolph Cowen, and Joe Healey to the Child Mind Institute. The views and opinions expressed in this article are those of the authors and should not be construed to represent the views of any of the sponsoring organizations, agencies, or U.S. Government.

### Competing Interest Statement

The authors have declared no competing interest.

### Funding Statement

This work was supported in part by the Intramural Research Program of the National Institute of Mental Health (ZIAMH002953) and Morgan Stanley. Additionally, by philanthropic gifts from Phyllis Green, Randolph Cowen, and Joe Healey to the Child Mind Institute. The views and opinions expressed in this article are those of the authors and should not be construed to represent the views of any of the sponsoring organizations, agencies, or U.S. Government.

### Author Declarations

Data are available to researchers by registering for a data usage agreement (http://fcon_1000.projects.nitrc.org/indi/cmi_healthy_brain_network/Pheno_Access.html#DUA).

